# *E-cadherin* Gene Promoter Hyper-methylation in Blood Samples from Breast Cancer Cases

**DOI:** 10.1101/2021.03.30.21254647

**Authors:** Sadia Ajaz, Sani-e-Zehra Zaidi, Saleema Mehboob Ali, Aisha Siddiqa, Muhammad Ali Memon

## Abstract

In carcinomas, dissemination of cancer cells via blood or lymph circulation constitutes an early event. E-cadherin is a transmembrane calcium dependent adhesion protein. Cellular de-differentiation and plasticity, underlying metastasis, is attributed to the loss of function of *E-cadherin* (*cdh1*) gene. The loss of gene expression may arise from promoter hypermethylation, which has been reported in multiple cancers. In the present pilot project, sixty (60) blood samples were collected from the breast cancer patients at a tertiary care hospital in Karachi, Pakistan. DNA was isolated from the cells circulating in the peripheral blood of the participants. Promoter hypermethylation was investigated through sodium-bisulfite treatment of DNA followed by methylation-specific PCR. In 53.3% of the patients, *E-cadherin* gene promoter hypermethylation was observed. Promoter hypermethylation of *E-cadherin* has been reported in DNA isolated from the tissue specimen. However, to the best of our knowledge this is the first report of *E-cadherin* promoter hypermethylation in cells isolated from the peripheral blood of breast cancer patients from a geographically specific population. The results have important implications in tumour staging and selection of treatment regimens.

## Introduction

E-cadherin is a trans-membrane calcium-dependent adhesion glycoprotein. The protein expression is mainly restricted to epithelial cells (Bruner and Derksen, 2018). It is a critical protein at adherens junction, where it connects the epithelial cells to each other (Halbleib and Nelson, 2006). E-cadherin is a tumour suppressor (Berx, 1995; Guilford 1998; Perl 1998; and Derksen 2006). In cancers, the cadherins are frequently non-functional through gene inactivation and/or functional inhibition. Consequently, these play siginficant role in tumourigenesis and progression (Wong, 2017). E-cadherin is considered to be clinically causal in carcinomas, particularly breast and gastric cancers (Derksen, 2006).

The encoding gene for E-cadherin (*CDH1*) is mapped to chromosome 16q22.1. A recent CRISPR/Cas9 experiment demonstrated tumour development and progression of breast carcinoma sub-type when dual incativation of *CDH1* and *PTEN* genes was carried out, (Annunziato, 2016). Clinically, inactivating germ-line and somatic mutations as well as *CDH1* promoter hypermethylation have been shown in breast cancers (Berx, 1995, 1996; Masciari, 2007; Strathdee, 2002). The epigenetic silencing of *CDH1* has also been reported in some human breast cancer cell lines (Yang, 2001; Szyf, 2004). However, in case of primary breast cancers, information regarding aberrant DNA methylation is limited. The current data shows frequency variation ranging from 21% to 80% (Cardeira, 2006; Liu, 2016), depending upon tumour sub-type and stage. In the present study, we report the promoter hypermethylation analysis for epigenetic silencing of *CDH1* gene in the circulting cells, which were collected from peripheral blood samples of breast cancer patients.

## Methodology

### Patients

A total of 67, with 60 eligible, blood samples were collected from the breast cancer patients at the Atomic Energy Medical Centre (AEMC), Jinnah Postgraduate Medical Centre (JPMC), Karachi, Pakistan were collected between July 2016 and July 2018. The blood samples were collected in ACD coated vacutainers (BD Vacutainer^®^ BD Franklin Lakes NJ USA) and either processed immediately or stored at 4°C. The study was approved by the ethical review committees (ERCs) of the participating institutions: the independent ERC, International Center for Chemical and Biological Sciences (ICCBS), University of Karachi, Karachi, Pakistan [ICCBS/IEC-016-BS/HT-2016/Protocol/1.0], and the Atomic Energy Medical Centre (AEMC), Jinnah Postgraduate Medical Centre (JPMC), Karachi, Pakistan [Admin-3(257)/2016]. The inclusion criteria for patients was diagnosis of primary breast cancer.

#### 1.1.1 Tumour Information

The information regarding cancer including histological classification, tumour size, tumour grade, tumour stage, lymph node metastasis, and status of estrogen receptor (ER), progestrone receptor (PR), and human epidermal growth factor receptor-2 (HER-2) were obtained from the patients’ medical records, where available. The patients were interviewed for age, family history of cancer, gynae and obs history, and ethnicity. The patients’ information was documented in the form of a questionnaire.

#### 1.1.2 Bi-sulfite Treatment of DNA Samples

DNA samples were treated with sodium bisulfite following standard protocol (Herman et al., 1996) with slight adjustments. 1µg of DNA sample (total volume adjusted to 45µl with autoclaved deionized water) was taken in 1.5ml microcentrifuged tube. For alkaline denaturation, 5.7µl of 3M NaOH was added to each DNA samples. Samples were then incubated for 17 minutes in shaking waterbath at 37°C, 50RPM. After incubation, unmethylated cytosine were converted to uracil by adding 530µl of freshly prepared mixture of 16mM hydroquinone and 4M liquid sodium bisulfite solution (pH adjusted to 5 with 1M NaOH solution) to each DNA samples. A drop of mineral oil was added to each sample. Samples were incubated for 16-18 hours, in shaking waterbath.

Second day, DNA samples were purified using DNA clean-up kit (Bio Basic^©^ 2017 Bio Basic Inc., UK) following the instructions. To each purified DNA, 5.7µl of 3M NaOH was added, followed by incubation at 37°C for 17 minutes. After incubation, DNA was preciptated with 10M ammonium acetate (17µl) and ice-cold absolute ethanol (500µl). Samples were kept at - 20°C for precepitation.

The next day, samples were centrifuged at maximam speed (21,000rcf) for 21 minutes at 4°C. The supernatant was discarded and pellets were washed twice with 70% ethanol (500µl), followed by centrifugation at maximam speed for 21 minutes. Supernatant was discarded and tubes were air dried at room temperture. DNA samples were then resuspended in 35µl of autoclaved deioninzed water. The samples were incubated in shaking waterbath at 37°C for 10 minutes for resuspension. DNA samples were then kept at 4°C till PCR.

#### 1.1.3 Methylation Specific Polymerase Chain Reaction (MS-PCR)

Primer sequences for methylated and unmethylated PCR are listed in Table 1.

| S.no | Gene | Forward primer<br>5' to 3' | Reverse primer<br>5' to 3' | Annealing<br>Temperature<br>(°C) | Product<br>size<br>(bp) | Reference |
| --- | --- | --- | --- | --- | --- | --- |
| 1. | *E-cadherin* | TAATTTTAGGT | CACAACCAAT | 54 | 97 |  |
|  | **UnMethylated** | TAGAGGGTTA<br>TTGT | CAACAACAC<br>A |  |  | Herman,<br>Graff, |
| 2. | *E-cadherin* | TTAGGTTAGA | TAACTAAAAA | 53.5 | 116 | Myöhänen |
|  | **Methylated** | GGGTTATCGC | TTCACCTACC |  |  | , Nelkin, |
|  |  | GT | GAC |  |  | and Baylin |
|  |  |  |  |  |  | (1996) |

PCR reactions for methylated and unmethylated genes were carried out separately.

For unmethylated PCR, reaction was carried out in a total volume of 25µl containing 1X PCR, 4.5mM MgCl_2_, 1.5mM dNTPs, 1.9µM/µl of primers, 3% DMSO, and 120ng of DNA.

Hot-start PCR was carried out. The samples were placed at 95°C for 5 minutes before the addition of 1.5U/µl Taq polymerase (Thermofisher scientific.Inc., USA). Amplification was carried out using Kyratec SC300 super cycler. PCR conditions were: initial denaturation at 95°C for 5 minutes followed by 35 cycles of 95°C for 30 seconds, 54°C for 30 seconds, 72°C for 30 seconds. Final extension was carried out at 72°C for 10 minutes (Herman et al., 1996).

For methylated PCR, PCR mix of 25µl comprised 1X PCR buffer, 6.7mM MgCl_2_ 1.5mM dNTPs, 1.85µM/µl of each primer, 10mM β-mercaptoethanol, 2U/µl Taq polymerase and 120ng of DNA. Cycling conditions were similar to unmethylated PCR. Anealing tempertaure was 53.5°C.

## Results

The clinico-pathological features of the patients’ samples included in the study are listed in Table 2.

**Table-2.** Clinicopathological characteristics of breast cancers cohort.

|  | **Parameter** | **N or value %** |
| --- | --- | --- |
| 1 | **Age (n\*=63)** |  |
| 1a | ≤40 | 26 (41.3%) |
| 1b | >40 | 37 (58.7%) |
| 2 | **Tumor Site (n\*=63)** |  |
| 2a | Right | 27 (42.2%) |
| 2b | Left | 35 (54.7%) |
| 2c | Bilateral | 01 (1.60%) |
| 3 | **Molecular Sub-type (n\*=35)** |  |
| 3a | Luminal A | 17 (48.6%) |
| 3b | Luminal B | 05 (14.3%) |
| 3c | Her 2/neu | 04 (11.4%) |
| 3d | Basal like | 09 (25.7%) |
| 4 | **Histological Sub-type (n\*=63)** |  |
| 4a | IDC | 54 (85.7%) |
| 4b | DCIS | 01(1.6%) |
| 4e | IDC + DCIS | 05 (7.9%) |
| 4f | Others | 03 (4.7%) |
| 5 | **Receptor Status (n\*=49 for ER & PR; n\*=37 for Her2-neu)** |  |
| 5a | ER positive | 31 (63.2%) |
| 5b | ER negative | 18 (36.7%) |
| 5d | PR positive | 30 (61.2%) |
| 5e | PR negative | 19 (38.8%) |
| 5g | Her2-neu positive | 10 (27%) |
| 5h | Her2-neu negative | 27 (73%) |
| 6 | **Tumor Size (n\*=58)** |  |
| 6a | T1(<2cm) | 09 (15.5%) |
| 6b | T2(2-5cm) | 37 (63.8%) |
| 6c | T3(>5cm) | 12 (20.7%) |
| 7 | **Stage (n\*=43)** |  |
| 7a | Low (I-II) | 21 (48.8%) |
| 7b | High (III-IV) | 22 (51.2%) |

“n\*”: number of samples with available data

Representative gels for *CDH1* methylation and un-methylation are shown in figures 1 and 2, respectively

**Figure 1.**
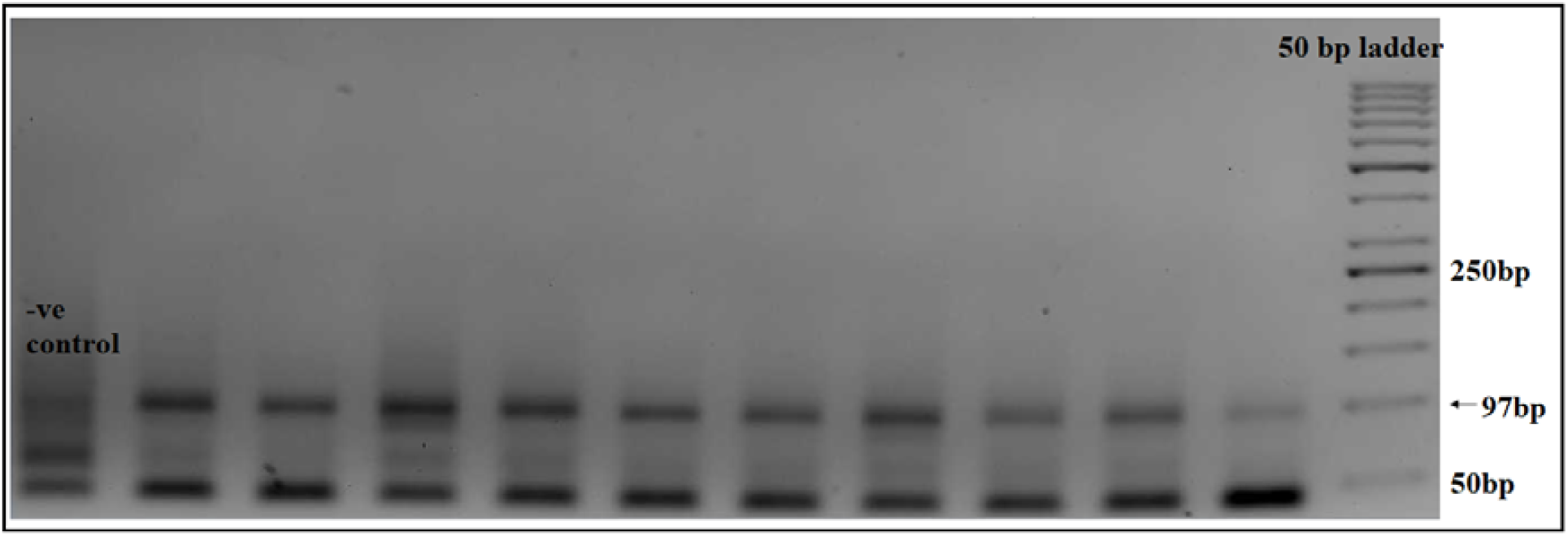
Agarose gel electrophoresis of MS-PCR product of UnMethylated *E-cadherin* promoter region, stained with ethidium bromide. Product size is 97bp.

**Figure 2.**
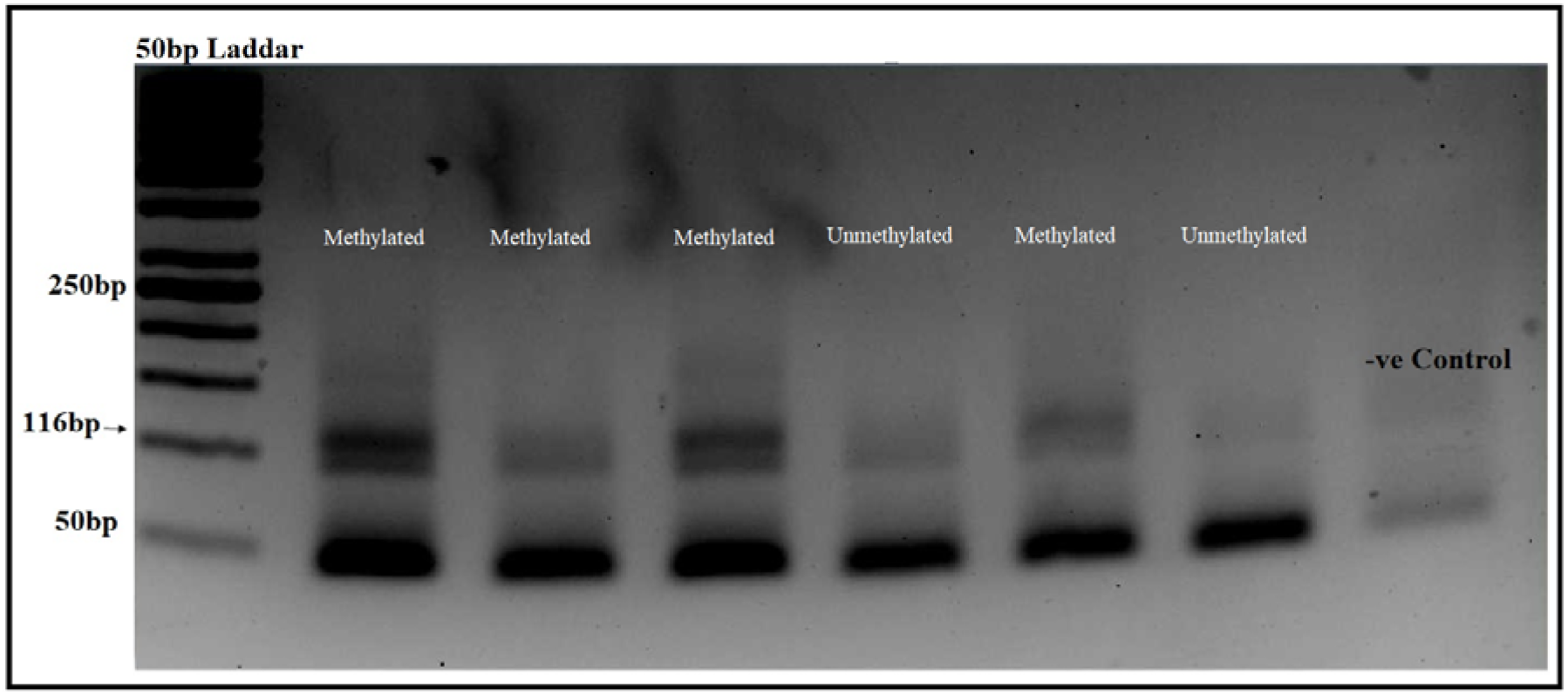
Agarose gel electrophoresis of MS-PCR product of methylated *E-cadherin* promoter region, stained with ethidium bromide. Product size is 116bp.

In the studied cohort of 60 breast cancer patients, *E-cadherin* hypermethylation was detected in 53.3% (32/60) of the cases. (Table 3)

**Table 3.** Methylation status of selected breast cancer patients.

| Methylation status | N=60* | Percentage (%) |
| --- | --- | --- |
| UnMethylated | 28 | 46.60% |
| Methylated | 32 | 53.3% |

\*Re

49% (18/36) of the cases are above 40 years while 58% (14/24) of the cases are below 40 years.

## Discussion

‘Epigenetics’ is combination of ‘epigenesis’ and ‘genomic’ which means ‘on top or in addition to genetics’. The term was coined six decades ago based on mechanisms of cell fate commitment and lineage specification (Holliday, 1990; Waddington, 1959). Epigenetic changes can reversibly inactivate tumor suppressor gene in a heritable manner (Llinas-Arias and Esteller, 2017). These gene(s) are either packed into heterochromatin or hypermethylation of CpG island occurs. Such changes result in gene silencing that may attribute to tumor progression (Alberts, 2017).

*E-cadherin* being cell adhesion molecule, plays significant role in cancer metastasis. Reduced expression of *E-cadherin* is one of the molecular events in cancer progression (Nass et al., 2000). In the studied cohort, hypermethylation is correlated with breast cancers progression and metastasis.

In the studied cohort comprising both low and advanced stage breast cancer patients, 53.3% of the cases showed promoter hypermethylation of *E-cadherin* in DNA from circulating cells. To the best of our knowledge this is the first report of *E-cadherin* promoter hypermethylation in cells isolated from the peripheral blood of breast cancer patients.

Two studies from Asia have analyzed *CDH1* promoter hyper-methylation in tissue samples from breast cancer patients. A study based on Kashmiri population has reported hypermethylation in 57.8% of the cases (Asiaf et al., 2014), whereas, another study from Iran reported hypermethylation in 94% of selected ductal-type breast cancer (Shargh et al., 2014).

In about 51% of IDC cases, *E-cadherin* hypermethylation was observed while in different studies, hypermethylation was observed in 30% of IDC cases (Nass et al., 2000).

## Conclusion

The pilot study concludes that *E*-*cadherin* hypermethylation is involved in breast cancer progression in our population. Further investigation should be conducted to evaluate the role of *E-cadherin* in cancer metastasis and response to treatment.

This work was presented in a preliminary form in an MPhil thesis of one of the co-authors (SZZ).

## Supporting information

STARD_CHECKLIST

## Data Availability

Data available on reasonable request.

